# Parents’ perspectives on a national child oral health promotion program: sociodemographic influences and behavioral insights

**DOI:** 10.1101/2025.09.25.25336673

**Authors:** Mohammad Reza Khami, Mohammadreza Naderi, Shabnam Varmazyari

## Abstract

**Background:** Iran’s *Students’ Oral Health Promotion Program* (SOHPP) aimed to improve primary school children’s oral health, but parental perceptions of this program, as key stakeholders, remain underexplored. This study explores parents’ perceptions of Iran’s SOHPP, the sociodemographic factors shaping them, and children’s post-program oral health behaviors.

**Methods:** Conducted at four randomly selected comprehensive healthcare centers in Tehran (July–August 2020), this cross-sectional study phone-surveyed parents of primary school children who participated in Iran’s SOHPP. The questionnaire covered sociodemographics, children’s post-program oral health behaviors, and awareness and satisfaction with key SOHPP components: oral health education, fluoride therapy, electronic oral health profiling, and treatment need identification. ANOVA, chi-square, and backward regression models served for statistical analysis.

**Results:** The 354 surveyed parents (response rate: 67%), on average, scored 79% for SOHPP awareness and 74% for satisfaction. Awareness and satisfaction were lowest for treatment-related components (58.2% and 52.0% for oral health profiling; 70.9% and 53.7% for treatment need identification). Fluoride therapy acceptance was 76.6%, with refusals mainly due to poor notification and limited procedural understanding. While 61.6% of parents noted improved tooth-brushing in their child, post-program, only 38.7% reported twice-daily brushing, and 37.9% were unaware of their fluoride toothpaste use. Additionally, 41.5% reported sugary snacking at least once daily by their child, while 83.0% reported healthy school food intake. More educated fathers had greater program awareness (B = 0.18, p = 0.040), satisfaction (B = 0.17, p = 0.032), and fluoride therapy acceptance (OR = 1.37, p = 0.024), whereas government-employed household heads were less aware (B = –1.16, p = 0.004) and less likely to perceive tooth-brushing improvements (B = –1.48, p = 0.001).

**Conclusions:** Enhanced treatment-related component delivery, improved fluoride therapy transparency, consistent oral health behavior promotion, and tailored outreach to parental sociodemographics are essential for improving how parents view Iran’s SOHPP.

## Introduction

Childhood oral health is a major global concern, with dental caries, the most common oral disease, affecting 60–90% of school-aged children worldwide (1, 2). Alongside periodontal disease, malocclusion, trauma, and noma, these conditions cause pain, impair daily functioning, disrupt learning and socialization, and have lasting effects into adulthood (3–5). The burden of these conditions is greatest in low- and middle-income countries (LMICs), where limited resources, poor access to care, and greater exposure to risk factors prevail (6, 7). In Iran, the situation is similarly alarming, with national data revealing high levels of untreated decay, few caries-free children, and worsening permanent teeth deterioration and periodontal problems as they age (8, 9). Despite substantial dental treatment needs among Iranian children, weak legislation, resource constraints, and inequitable service distribution hinder them enjoying optimal oral health (9–13).

In response to the global oral health crisis among children, child oral health promotion programs have received increasing attention (14). These programs improve clinical oral health outcomes by reducing dental caries and plaque, enhance oral health knowledge, attitudes, and behaviors, expand equitable access to preventive services, and generate cost savings for health systems (14–18). Examples include Kuwait’s national School Oral Health Program, which combines education, prevention, and treatment through fluoride varnish, sealants, restorative care, and supervised brushing, and South Korea’s children’s dental care program, which provides free annual registration with dentists for preventive and restorative services (18, 19).

In Iran, the *Students’ Oral Health Promotion Program* (SOHPP) represents a similar effort. Launched in 2015 as part of Iran’s Ministry of Health’s Oral Healthcare, the SOHPP focused specifically on the country’s nine million primary school students through integrated oral health education, prevention, and treatment efforts. Oral health education involved dentists training school teachers and healthcare providers at comprehensive health centers, Iran’s primary healthcare hubs (9), to in turn educate children and parents using workshops, demonstrations, and audiovisual aids. Prevention centered on twice a year fluoride varnish therapy at schools, while treatment included school-based oral cavity screenings, electronic oral health profile establishment, and referrals for care to comprehensive healthcare centers, charities, and private providers (9, 20).

Meaningful stakeholder engagement is essential for community-based dental programs such as Iran’s SOHPP to ensure their relevance, acceptability, and sustainability (21–23). Regular stakeholder evaluations, as a key component of such engagement, further strengthens program effectiveness and guides authorities on whether to sustain, adapt, or discontinue these programs (24). Parents are key stakeholders in children’s oral health (25). They impact children’s oral health knowledge, attitudes, and practices, affect their clinical oral health outcomes, and shape their long-term oral health trajectories through their own sociodemographic characteristics (21, 26–28). Yet, research on parental perceptions of child oral health promotion programs remains limited. Only a few studies, including Qatar’s *Asnani* and Australia’s *Bright Smiles Bright Futures* have addressed this issue (29, 30), while most research has concentrated on child oral health outcomes following program implementation (21, 31). This gap is also seen in Iran’s SOHPP, where evaluations have largely overlooked parents’ perspectives and emphasized clinical outcomes, such as dental caries experience (24, 32).

Therefore, for the first time to the best of our knowledge, this study aims to evaluate Iranian parents’ perspectives on the country’s SOHPP, identify the sociodemographic factors shaping these perspectives, and explore parents’ perceptions of their children’s oral health behaviors following program implementation. By doing so, it hopes to generate evidence for improving SOHPP’s design, tailoring its implementation, and ensuring its equitable uptake across diverse family contexts.

## Methods

### Study design and ethical considerations

The cross-sectional analytical study was conducted between July 1^st^ and August 31^st^ 2020, after obtaining approval from the Ethics Committee of Tehran University of Medical Sciences (TUMS) (ethics code: IR.TUMS.DENTISTRY.REC.1399.011). Participation in the study was voluntary, and verbal informed consent was obtained from the participants after thoroughly explaining to them the voluntary nature of participation, their right to withdraw participation at any time, and the efforts exerted for maintaining confidentiality and anonymity. These efforts included obtaining phone numbers from the Health and Treatment Deputies of TUMS without any accompanying personally identifiable information and filling out all questionnaires anonymously.

### Sample Size calculation

Since no sufficiently similar prior research was found, the main sample size was estimated using a pilot study on 30 parents. Based on the pilot study and using the one-sample proportion confidence interval function in SPSS (version 26), the minimum required sample size was calculated as 347, assuming α = 0.05, p = 0.69, and d = 0.1.

### Study setting, population, and procedure

The study was conducted at four randomly selected Comprehensive Healthcare Centers within TUMS jurisdiction in Tehran city, Iran. Iran’s healthcare system is overseen by its medical universities, and in Tehran, the southern region falls under TUMS’s jurisdiction (33). Tehran city was chosen for its logistical feasibility, large population, and status as the nation’s capital, while the TUMS jurisdiction was selected for its high population density and accessibility (34, 35).

Parents were eligible to participate if they had a child in grades 1–6 that had participated in the SOHPP, were registered at one of the selected Comprehensive Healthcare Centers under TUMS jurisdiction in Tehran city, had a reachable phone number on record, and provided informed consent.

Data for this study were collected using a structured questionnaire. The research team first obtained the necessary approvals from TUMS and requested a list of comprehensive healthcare centers under their supervision. Four centers, *Ayat, Farmafarmanian, Imam Hassan Mojtaba, and Avicenna*, were randomly selected from this list. The research team visited these centers to obtain parent contact information. Then, one of the researchers conducted telephone interviews with parents using a questionnaire developed for the study.

### Study tool and measures

Data were collected using a Persian questionnaire developed for this study (supporting file 1). This questionnaire was checklist-based in nature, did not evaluate complex constructs (such as oral health knowledge or attitudes) across multiple domains or with Likert scales, and merely captured whether parents were aware of, satisfied with, and accepting of specific components of the SOHPP. In addition, only one member of the research team administered this questionnaire via telephone interviews, who was actively available to explain each item, clarify response options, and ensure accurate understanding throughout the call. Therefore, simplified psychometric validation procedures were deemed appropriate. Face and content validity were assessed by discussions among faculty experts in Community Oral Health through group, and reliability was evaluated using a test-retest method with a subsample of 20 parents. Feedback from these procedures was used to revise questionnaire where necessary.

The instrument included four main sections that asked about: 1) parents’ sociodemographic characteristics, 2) their awareness and satisfaction with the four key SOHPP components (oral health education, fluoride therapy, electronic oral health profiles, and treatment need identification), 3) their acceptance/perceived impact of certain program components, and 4) their reports on post-program oral health behaviors of their child. More details on the items in these sections, their response options, and how they were grouped to form the research measures are provided below:

Sociodemographic characteristics: This section included items on the respondent’s relation to the child (mother/father/other), father’s level of education (illiterate, literate, primary school, middle school, high school diploma, associate degree, bachelor’s degree, master’s degree, doctorate or higher, do not know), mother’s level of education (similar items); household size (in discrete numbers), household head occupation (government employee, government sector worker, private sector employee, private sector worker, self-employed, homemaker, retired, unemployed), and the child’s school grade (numbers between 1 to 6). These characteristics were treated as independent variables in analyses.

The derived outcome variables included:

a. Overall awareness of SOHPP: Defined as having awareness that SOHPP exists and offers specific services, did not imply approval or participation. This factor was assessed using four items covering oral health education, fluoride therapy, electronic oral health profiling, and identification of treatment needs. Responses were coded as Yes = 2, No opinion = 1, No = 0. A composite awareness score ranging from 0–8 was calculated by summing scores for these 4 items, with higher scores indicating greater awareness.
b. Overall satisfaction with SOHPP: Defined as perceived service quality and subjective evaluation of program delivery, not behavioral compliance or endorsement. This factor was assessed using four items corresponding to the same above noted four program components. Responses followed the same coding scheme as the awareness variable. A composite satisfaction score ranging from 0–8 was calculated by summing scores for these 4 items, with higher scores reflecting greater satisfaction
c. Acceptance of fluoride therapy: Defined as behavioral consent to the intervention. This factor was assessed through a single item asking whether parents agreed with fluoride application for their children. Response options included “Yes”, “No opinion”, and “Other”, with the parents stating their reason for refusal if opting for the last option.
d. Perceived impact of SOHPP on child’s tooth-brushing habits: Defined as parent’s belief that the SOHPP had influenced their child’s tooth-brushing behavior, reflecting perceived behavioral change, not objective behavior. This factor was assessed through a single item asking whether the parent thought the program had influenced their child’s tooth-brushing behavior. Response options included “Positive”, “No opinion”, and “Negative”.
e. Daily tooth-brushing frequency: This factor was assessed through a single item asking how often the child brushed their teeth per day post-program implementation. Response options included “At least twice a day”, “Once a day”, “Less than once a day”, and “Never”.
f. Use of fluoride toothpaste: This factor was assessed through a single item asking if the child used fluoride toothpaste during tooth-brushing post-program implementation. Response options included “Yes”, “No”, and “No opinion”.
g. Frequency of sugary snack consumption: This factor was assessed through a single item asking how frequently the child consumed sugary snacks post-program implementation. Response options included “At least once a day”, “Less than once a day”, and “Never”.
h. Type of foods consumed at school: This factor was assessed through a single open-ended item asking what types of foods the child typically ate at school post-program implementation. One of the researchers reviewed the responses and classified them according to the World Health Organization’s (WHO) nutritional criteria (33), which defines healthy foods as those low in added sugars, saturated fats, and sodium and unhealthy foods as those high in added sugars, saturated fats, or sodium. For example, responses listing “milk,” “bread with cheese,” or “fruits” were coded as “Healthy,” while “chips,” “chocolate,” or “sugar-sweetened drinks” were coded as “Unhealthy.”

### Statistical Analysis

Data were analyzed using SPSS version 26.0. Descriptive statistics including means, standard deviations, frequencies, and percentages were computed to summarize demographic characteristics and key variables related to parental awareness, satisfaction, and behavioral outcomes. Group comparisons were conducted using ANOVA, Chi-square tests, and independent sample t-tests where appropriate.

To examine associations between independent and outcome variables, multiple linear regression analyses were conducted for continuous outcomes (overall awareness, overall satisfaction, and perceived impact on tooth-brushing), while binary logistic regression was used for the dichotomous outcome (acceptance of fluoride therapy). A backward elimination method was applied in each regression model, whereby predictors were progressively removed if they did not meet the 0.10 threshold. Final models retained only predictors with p-values less than 0.05.

Model fit, assumptions of normality, linearity, and homoscedasticity were evaluated. Multicollinearity was assessed using variance inflation factors (VIFs), all of which were below 5, indicating acceptable levels. Occupation, a nominal variable, was dichotomized for inclusion in the regression models. Categories with small sample sizes, such as unemployed and retired individuals, had to be excluded. Government employees were coded as 0, while all other occupations were coded as 1, reflecting the considerable differences observed in scores between these groups. Statistical significance was set at p < 0.05 for all tests and missing data were handled using listwise deletion.

## Results

Of the 525 individuals approached, 354 participated, yielding a 67% response rate, with an average of 87 participants per comprehensive healthcare center. Fathers comprised the largest group of respondents (47.2%). Among fathers, those with a college degree or higher were most common (30.9%), and those with primary education or less were least common (12.9%). For mothers, high school graduates predominated (28.2%), with primary education or less being the least common (19.2%). Regarding household head occupation, nearly half of the sample were self-employed (47.5%), while retirees were the smallest group (1.7%). About two-fifths of the households had four members (42.9%), and those with seven or more members were rare (2%). Overall, parents of children in grades 1–4 represented nearly 90% of the sample.

The mean parental awareness score was 6.33 (SD = 1.97), corresponding to 79% of the maximum score. Overall, 71.6% of parents scored 8, indicating full knowledge of SOHPP implementation, while 13.4% scored 0, reflecting complete unawareness. Awareness varied by program component, from 80.8% for oral health education to 58.2% for electronic oral health profile establishment (Table 2). The mean satisfaction score was 5.90 (SD = 1.72), or 74% of the maximum, with 58.8% of parents scoring 8 (complete satisfaction) and 11.2% scoring 0 (complete dissatisfaction). Satisfaction also varied by program component, from 68.6% for oral health education to 52% for electronic profile establishment. Additionally, 61.6% of parents perceived a positive impact of SOHPP on their child’s toothbrushing habits, while 76.6% accepted fluoride therapy and 15.8% declined. Parents’ reasons for refusing school-based fluoride therapy are ranked below by frequency of citation:

1. Lack of prior notification from schools about the intervention
2. Limited knowledge about fluoride therapy procedures
3. Concerns of improper provision in school settings
4. Belief that fluoride therapy increases susceptibility to dental caries
5. Fear of side effects such as gum or tongue redness and burning
6. Preference for a dental office
7. Child-specific issues such as psychological or physical conditions
8. Uncertainty regarding whether or not consent was required for the procedure

**Table 1.** Sociodemographic characteristics of parents of Tehran’s primary school children participating in the SOHPP^a^ (n = 354)

| Variables | Category | Number (%) |
| --- | --- | --- |
| Father's education level | Primary education or lower | 46 (12.9%) |
|  | Middle school education | 101 (28.5%) |
|  | High school diploma | 98 (27.7%) |
|  | Associate degree or higher | 109 (30.9%) |
| Mother's education level | Primary education or lower | 68 (19.2%) |
|  | Middle school education | 91 (25.7%) |
|  | High school diploma | 100 (28.2%) |
|  | Associate degree or higher | 95 (26.9%) |
| Household head's occupation | Self-employed | 168 (47.5%) |
|  | Private-sector laborer | 60 (16.9%) |
|  | Private-sector employee | 52 (14.7%) |
|  | Government laborer | 31 (8.8%) |
|  | Government employee | 25 (7.1%) |
|  | Unemployed | 12 (3.4%) |
|  | Retiree | 6 (1.7%) |
| Household size | 3 | 90 (25.4%) |
|  | 4 | 152 (42.9%) |
|  | 5 | 64 (18.1%) |
|  | 6 | 41 (11.6%) |
|  | 7 or higher | 7 (2%) |
| Relation of respondent to the child | Mother | 146 (41.3%) |
|  | Father | 169 (47.7%) |
|  | Other | 39 (11%) |
| Child's school grade | First | 82 (23.2%) |
|  | Second | 83 (23.4%) |
|  | Third | 87 (24.6%) |
|  | Fourth | 69 (19.5%) |
|  | Fifth | 28 (7.9%) |
|  | Sixth | 5 (1.4%) |

<sup>a</sup>SOHPP: Students' oral health promotion program

**Table 2.** Perceptions of SOHPP^a^ components among parents of Tehran’s primary school children (n = 354)

| Variable | Category | N (%) |
| --- | --- | --- |
| Awareness of electronic oral health profile establishment | No | 72 (20.3%) |
|  | No opinion | 76 (21.5%) |
|  | Yes | 206 (58.2%) |
| Awareness of fluoride therapy provision | No | 30 (8.5%) |
|  | No opinion | 53 (15.0%) |
|  | Yes | 271 (76.6%) |
| Awareness of oral health education | No | 29 (8.2%) |
|  | No opinion | 39 (11.0%) |
|  | Yes | 286 (80.8%) |
| Awareness of treatment need identification | No | 59 (16.7%) |
|  | No opinion | 44 (12.4%) |
|  | Yes | 251 (70.9%) |
| Satisfaction with electronic oral health profile establishment | No | 15 (4.2%) |
|  | No opinion | 155 (43.8%) |
|  | Yes | 184 (52.0%) |
| Satisfaction with fluoride therapy provision | No | 59 (16.7%) |
|  | No opinion | 79 (22.3%) |
|  | Yes | 216 (61.0%) |
| Satisfaction with oral health education | No | 43 (12.1%) |
|  | No opinion | 68 (19.2%) |
|  | Yes | 243 (68.6%) |
| Satisfaction with treatment need identification | No | 42 (11.9%) |
|  | No opinion | 122 (34.5%) |
|  | Yes | 190 (53.7%) |
| Fluoride therapy acceptance | Yes | 271 (76.6%) |
|  | No | 56 (15.8%) |
|  | Other | 27 (7.6%) |
| Perceived Impact on tooth-brushing habit | Negative | 84 (23.7%) |
|  | No opinion | 52 (14.7%) |
|  | Positive | 218 (61.6%) |

<sup>a</sup>SOHPP: Students' oral health promotion program

**Table 3.**
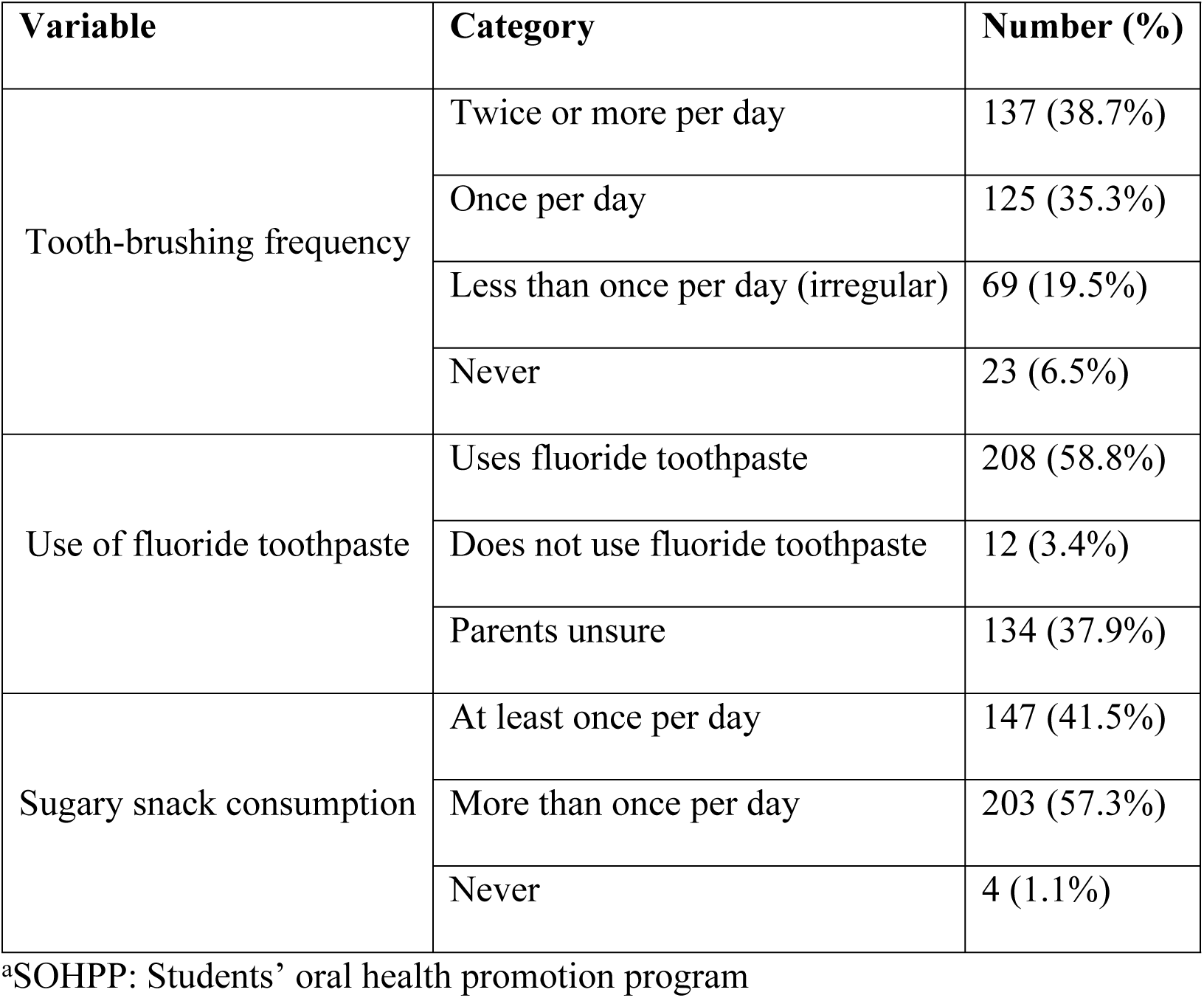
Parental reports of the oral health behaviors of Tehran’s primary school children post-SOHPP^a^ (n=354)

Approximately two-fifths of parents (38.7%) reported that their child brushed at least twice daily after the SOHPP. Nearly three-fifths (58.8%) said their child used fluoride toothpaste, although about two-fifths (37.9%) were unsure of its use. Meanwhile, 41.5% indicated that their child consumed sugary snacks at least once a day, whereas a large majority (83%) reported that their child ate healthy foods at school. The healthy items mentioned were milk, bread with cheese, fruits and vegetables, and nuts, while unhealthy items ones were dried sour plums, chips, puffed snacks, chocolate, chewing gum, and sugar-sweetened beverages.

One-way ANOVA indicated significant differences in mean overall satisfaction scores across levels of fathers’ education (p = .011). Post-hoc comparisons revealed that fathers with primary education or lower reported lower satisfaction compared to those with high school diplomas or associate degrees or higher. Similarly, there were significant variations in overall awareness scores by fathers’ education (p < .001), with post-hoc analyses indicating progressively higher awareness among those with middle school education, high school diplomas and associate degrees or higher. For mothers’ education, only awareness scores differed (p = .018), driven by higher means among those with high school diplomas compared to primary education or lower. Household head occupation was associated with significant differences in awareness (p = .007), with unemployed heads reporting the lowest scores (mean = 4.58, SD = 2.2), though satisfaction did not vary significantly (p = .401) (Table 4).

**Table 4.** SOHPP^a^ Awareness and Satisfaction by Sociodemographics among Parents of Tehran’s Primary School Children (n=354)

| Variable | Category | Overall SOHPP Awareness |  |  | Overall SOHPP Satisfaction |  |  |
| --- | --- | --- | --- | --- | --- | --- | --- |
|  |  | Mean | SD | P-value | Mean | SD | P-value |
| Father's education level | Primary education or lower | 5.2 | 2.5 | <.001* | 5.1 | 2.2 | .011* |
|  | Middle school education | 6.3 | 1.7 |  | 5.9 | 1.7 |  |
|  | High school diploma | 6.5 | 1.7 |  | 6.1 | 1.5 |  |
|  | Associate degree or higher | 6.6 | 2.0 |  | 6.0 | 1.6 |  |
| Mother's education level | Primary education or lower | 6.0 | 2.1 | .018* | 5.8 | 2.0 | .766 |
|  | Middle school education | 6.0 | 2.0 |  | 6.0 | 1.5 |  |
|  | High school diploma | 6.8 | 1.7 |  | 6.0 | 1.7 |  |
|  | Associate degree or higher | 6.4 | 2.0 |  | 5.8 | 1.7 |  |
| Household head's occupation | Self-employed | 6.4 | 2.0 | .007* | 5.9 | 1.7 | .401 |
|  | Private-sector laborer | 6.5 | 1.5 |  | 6.0 | 1.9 |  |
|  | Private-sector employee | 6.4 | 2.0 |  | 5.9 | 1.5 |  |
|  | Government laborer | 6.3 | 2.1 |  | 6.1 | 1.9 |  |
|  | Government employee | 5.4 | 2.3 |  | 5.4 | 1.6 |  |
|  | Unemployed | 4.6 | 2.2 |  | 5.5 | 1.7 |  |
|  | Retiree | 7.3 | 0.8 |  | 7.0 | 0.6 |  |
| Household size | 3 | 6.4 | 1.9 |  | 5.8 | 1.7 |  |
|  | 4 | 6.4 | 1.6 |  | 6.0 | 1.8 |  |
|  | 5 | 6.3 | 2.3 | .196 | 5.9 | 1.5 | .985 |
|  | 6 | 5.7 | 2.5 |  | 5.9 | 1.7 |  |
|  | 7 or higher | 5.7 | 1.7 |  | 5.7 | 1.7 |  |
| Relation to the child | Mother | 6.4 | 2.0 |  | 5.9 | 1.7 |  |
|  | Father | 6.4 | 1.8 | .120 | 5.9 | 1.7 | .719 |
|  | Other | 5.7 | 2.4 |  | 6.0 | 1.9 |  |
| Child's school grade | First | 6.3 | 2.0 |  | 5.7 | 1.8 |  |
|  | Second | 6.2 | 2.0 |  | 5.9 | 1.8 |  |
|  | Third | 6.4 | 2.0 | .820 | 6.0 | 1.7 | .544 |
|  | Fourth | 6.5 | 1.8 |  | 6.1 | 1.5 |  |
|  | Fifth | 6.0 | 2.2 |  | 5.6 | 1.8 |  |
|  | Sixth | 6.4 | 0.9 |  | 5.6 | 2.3 |  |

<sup>a</sup>SOHPP: Students' oral health promotion program; \*P-values below 0.05 were deemed
statistically significant.

As shown in Table 5, higher father education was associated with greater overall awareness of the program (B = 0.18, p = 0.040), higher overall satisfaction (B = 0.17, p = 0.032), and greater approval of the child receiving fluoride therapy (OR= 1.37, p = 0.024). In contrast, being a government employee as a household head was related to lower overall awareness of the program (B = –1.16, p = 0.004) and less likelihood of deeming the program had had a positive impact on child tooth-brushing frequency (B = –1.48, p = 0.001).

**Table 5.**
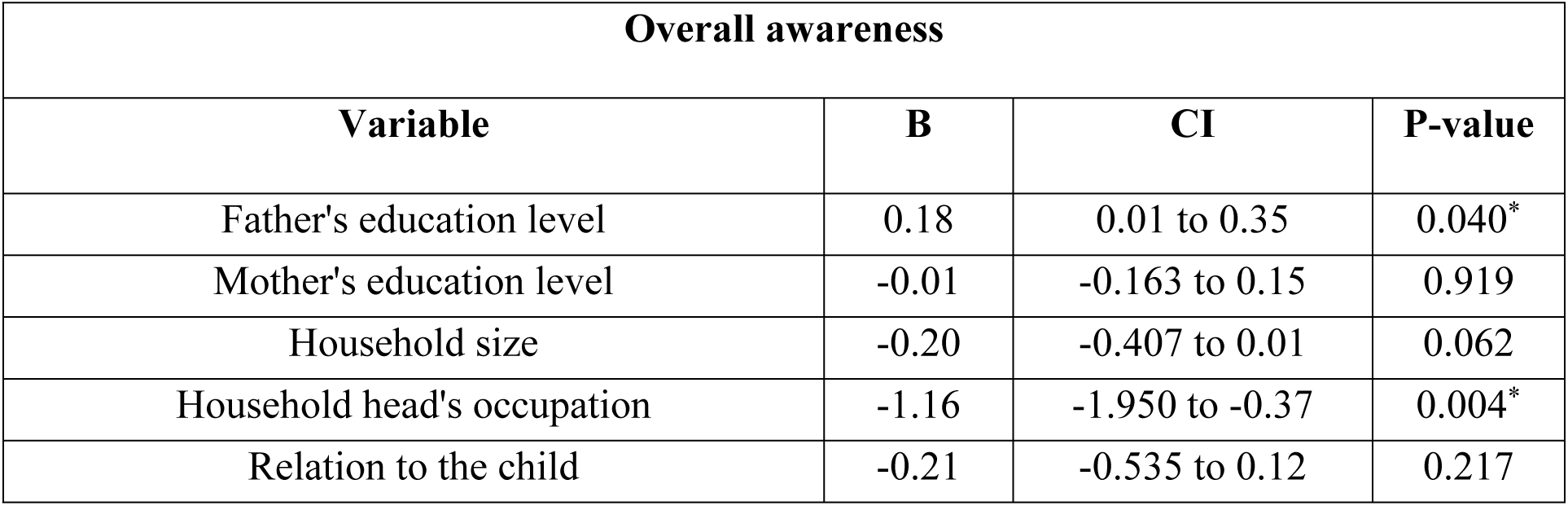
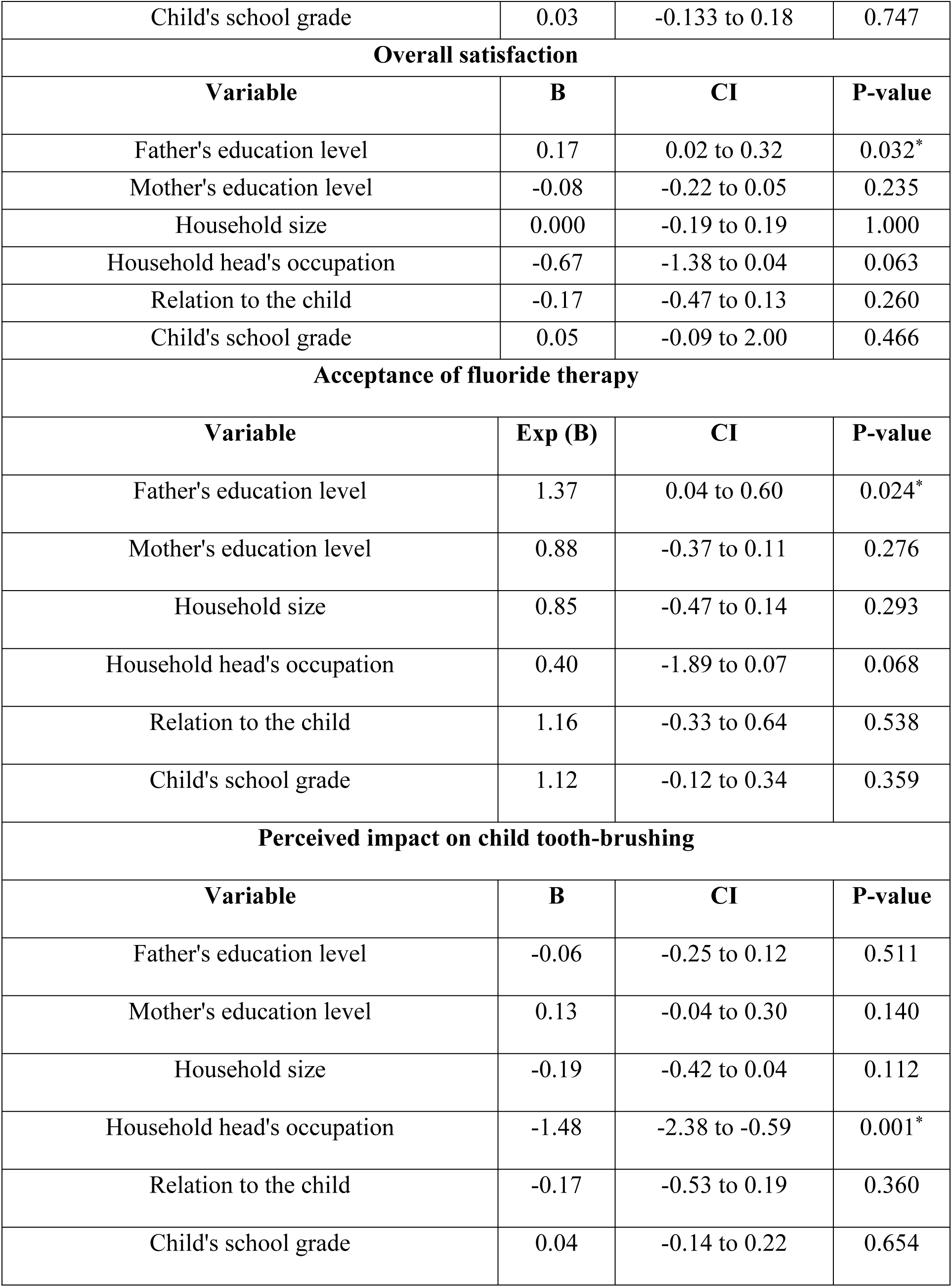
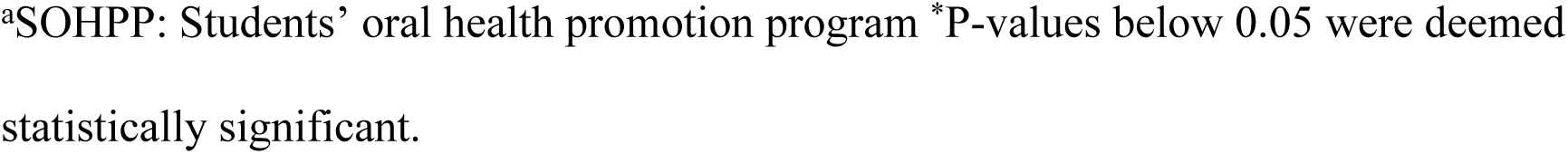
Sociodemographic Indicators of SOHPP^a^ perceptions among parents of Tehran’s primary school children (n=354)

| Overall awareness |  |  |  |
| --- | --- | --- | --- |
| Variable | B | CI | P-value |
| Father's education level | 0.18 | 0.01 to 0.35 | 0.040* |
| Mother's education level | -0.01 | -0.163 to 0.15 | 0.919 |
| Household size | -0.20 | -0.407 to 0.01 | 0.062 |
| Household head's occupation | -1.16 | -1.950 to -0.37 | 0.004* |
| Relation to the child | -0.21 | -0.535 to 0.12 | 0.217 |
| Child's school grade | 0.03 | -0.133 to 0.18 | 0.747 |
| **Overall satisfaction** |  |  |  |
| **Variable** | **B** | **CI** | **P-value** |
| Father's education level | 0.17 | 0.02 to 0.32 | 0.032* |
| Mother's education level | -0.08 | -0.22 to 0.05 | 0.235 |
| Household size | 0.000 | -0.19 to 0.19 | 1.000 |
| Household head's occupation | -0.67 | -1.38 to 0.04 | 0.063 |
| Relation to the child | -0.17 | -0.47 to 0.13 | 0.260 |
| Child's school grade | 0.05 | -0.09 to 2.00 | 0.466 |
| **Acceptance of fluoride therapy** |  |  |  |
| **Variable** | **Exp (B)** | **CI** | **P-value** |
| Father's education level | 1.37 | 0.04 to 0.60 | 0.024* |
| Mother's education level | 0.88 | -0.37 to 0.11 | 0.276 |
| Household size | 0.85 | -0.47 to 0.14 | 0.293 |
| Household head's occupation | 0.40 | -1.89 to 0.07 | 0.068 |
| Relation to the child | 1.16 | -0.33 to 0.64 | 0.538 |
| Child's school grade | 1.12 | -0.12 to 0.34 | 0.359 |
| **Perceived impact on child tooth-brushing** |  |  |  |
| **Variable** | **B** | **CI** | **P-value** |
| Father's education level | -0.06 | -0.25 to 0.12 | 0.511 |
| Mother's education level | 0.13 | -0.04 to 0.30 | 0.140 |
| Household size | -0.19 | -0.42 to 0.04 | 0.112 |
| Household head's occupation | -1.48 | -2.38 to -0.59 | 0.001* |
| Relation to the child | -0.17 | -0.53 to 0.19 | 0.360 |
| Child's school grade | 0.04 | -0.14 to 0.22 | 0.654 |

<sup>a</sup>SOHPP: Students' oral health promotion program \*P-values below 0.05 were deemed statistically significant.

## Discussion

Iran’s SOHPP offers a noteworthy example of large-scale, national child oral health promotion program, integrating oral health education, prevention, and treatment into routine student health and leveraging intersectoral collaboration to reach diverse family dynamics. This study examined parents’ perceptions of Iran’s SOHPP, highlighting positive trends and remaining gaps while stressing the need for tailored strategies to improve parental views.

Parents in this study showed generally high awareness and satisfaction with the SOHPP, scoring on average 79% for awareness and 74% for satisfaction. Additionally, 61.6% felt the program had positively affected their child’s tooth brushing. Similar findings have been reported in other child oral health programs, such as Australia’s *Bright Smiles Bright Futures* and Qatar’s *Asnani* program, where parents expressed appreciation, gratitude, and active engagement (29, 30).

Notably, the Australian study highlighted also children’s growing independence and enthusiasm, while the Qatari study reported even stronger effects, with children actively reminding parents of porgam teachings and participating through school reinforcement and gamified tools (29, 30).

These programs were more interactive, gamified, supervised, and better followed up compared to Iran’s SOHPP, suggesting potential areas for improvement. In addition, parents were most aware of and satisfied with the oral health education and fluoride therapy components, but, less so with the treatment-related components of oral health profiling and treatment need identification. This may reflect these components’ less interactive and visible nature, and possibly unmet expectations if identified treatment needs were not followed-up and referred adequately, given the large student population and the complexity of organizing care.

Fluoride therapy acceptance in the present study was high, with 76.6% of parents accepting the intervention for their children. This high acceptance rate contrasts with lower rates reported in a U.S. study (58.7%) (36), but aligns with studies in Laos (80% for silver diamine fluoride after educational videos) and the UK and U.S. (approximately 70% for silver diamine fluoride in posterior teeth) (37, 38). In this study, most of the fluoride therapy refusals were due to logistical, procedural, or trust issues with school-based delivery, with safety concerns accounting for approximately one-third of responses. This differs from international findings, where fluoride therapy refusals were often due to fears of toxicity or developmental harm, low awareness, aesthetic concerns, personal choice, or media-fueled distrust (38–42). These differences may reflect the fact that this study examined parents’ perceptions of a child oral health promotion program, an aim not addressed in the other cited studies. In this context, improving procedural clarity and communication is likely to foster greater fluoride therapy acceptance than focusing strongly on safety or aesthetic concerns.

Although baseline data were not collected, parental reports of children’s oral health behaviors after program implementation still provide valuable information. According to parents, fewer than two-fifths of children brushed their teeth twice daily, while the rest brushed once a day (35.3%), less than once a day (19.5%), or not at all (6.5%), clearly falling short of the internationally recommended twice-daily guideline (43). This prevalence was lower than national averages for 6- and 12-year-olds (47.9% and 55.2%) (44), but considerably higher than the 9.3% reported in a prior Tehran study (45). This might indicate that the program had a positive local impact but did not meet national benchmarks. However, comparisons across these studies should be made cautiously, given their vast design, sampling, and measurement differences. Interestingly, despite the suboptimal prevalence reported for twice-daily tooth-brushing 61.6% of parents deemed that the SOHPP has positively impacted their child’s tooth-brushing, highlighting a perception–behavior gap. Similarly, although only 3.4% reported non-use of fluoride toothpaste, nearly 38% were unsure of their child’s usage. These findings suggest that parents’ optimism, potentially stemming from their notable awareness and satisfaction with the SOHPP, did not align with actual child oral health behaviors, and that greater emphasis on monitoring, reinforcement, and parental engagement is needed to translate perceived program benefits into sustained behaviors. Regarding diet, about two-fifths of children reportedly consumed sugary snacks at least once daily, though the survey did not capture more frequent intake, likely underestimating true consumption. Actual intake is likely higher, with prior estimates suggesting about eight daily servings among Iranian children (46). Thus, sugar exposure likely remained a behavioral risk post-SOHPP, despite the WHO’s recommendations to limit sugary snacking (47). Encouragingly, 83% of parents reported that the children mainly consumed healthy foods at school, suggesting the school environment reinforced the program’s nutritional efforts. Overall, while the SOHPP may have supported some positive oral health behaviors in its target population, notable gaps persist, emphasizing the need for continued interventions.

Despite the often underappreciated role of fathers in child oral health, fathers’ education emerged as a significant indicator of SOHPP perceptions in the present study, as higher-educated fathers demonstrated greater program awareness, higher program satisfaction, and stronger acceptance of fluoride therapy. This fits with evidence showing that higher education helps people find and use health information, is linked to higher socioeconomic status with better access to dental care and hygiene products, builds more trust in public health systems, and makes it easier to navigate healthcare services (48–53). Interestingly, maternal education did not significantly indicate SOHPP perceptions, contrasting with the existing literature which generally identifies mothers as primary agents of child oral health and maternal education as a key indicator of related oral health behaviors (27, 54). Sociocultural patterns in Iran and the broader Middle East may explain this discrepancy, as patriarchal norms in these regions often assign fathers primary authority in health decision-making, while mothers focus on daily caregiving tasks (55–57). Since traditional assumptions about maternal influence seem to not apply in all cultural contexts, local family dynamics should be considered when implementing child oral health programs.

Household heads employed in the government sector were less likely to be aware of the SOHPP or to perceive a positive impact on their child’s tooth brushing. Several factors may explain this. Structured, bureaucratic jobs may have limited opportunities to encounter program information, since demanding work can reduce parental engagement in child oral health due to less time and higher stress (58, 59). Additionally, awareness of public sector inefficiencies may have fostered skepticism, as perceptions of corruption or ineffectiveness can lower trust in government-led programs (60). Alternatively, these parents may already have higher oral health literacy and established tooth-brushing routines for their child, making the program seem less valuable, a notion partially supported by a study showing French government employees had higher oral health knowledge than the general population (61). These hypotheses warrant further investigation, particularly to guide targeted outreach for different subgroups of working parents.

Finally, it is worth noting that although household size did not reach statistical significance in relation to parents’ SOHPP awareness or perceived impact on child tooth-brushing, the negative coefficients suggested a possible attenuation effect. Larger households may face competing demands on time and resources that dilute health messaging (62, 63), though some studies suggest they benefit from shared caregiving and social support. (64–66). Given the mixed evidence and lack of statistical significance in the present findings, future research should explore how household composition relates to parental perceptions of child oral health promotion efforts.

Multi-level strategies are needed to enhance parents’ perspectives on Iran’s SOHPP. Improving delivery of treatment-related components, such as providing online or printed summaries of children’s oral health profiles and timely follow-up on treatment needs, could enhance awareness and satisfaction. High fluoride therapy acceptance provides a foundation to expand preventive care in the program, while refusals can be reduced through clear communication, prior notification, and procedural transparency. Consistency in children’s tooth-brushing habit can be promoted with logs, digital reminders, and gamified incentives, and healthier snacking both at home and at school can be encouraged through nutrition education sessions for parents and partnerships with school vendors. Parental sociodemographic factors should also be taken into consideration when implementing the program. These can include culturally appropriate, simplified information about the program for fathers with lower education and flexible outreach via workplaces, SMS, or digital platforms for government-employed household heads.

This study has several strengths, including its comprehensive data collection, a relatively strong response rate of 67% for a telephone-based survey, the use of random sampling, a validated questionnaire, and a focus on parental perspectives, which represents a critical gap in the literature. The interviewer-administered, telephone-based format was selected to enhance response accuracy compared with self-administered surveys and to enable safe data collection during the COVID-19 pandemic.

Several limitations should be acknowledged. The cross-sectional design, though suitable for the research aim, restricted causal inference. Reliance on self-reports raised risks of recall and social desirability bias, likely amplified by interviewer-led calls, while phone-based recruitment may have excluded households without stable phone access. The TUMS jurisdiction was chosen for feasibility during COVID-19, but Tehran’s greater socioeconomic resources and healthcare access compared to rural areas may have limited generalizability and inflated parental awareness, satisfaction, and child oral health behaviors. The absence of baseline data, consistent with the study’s aim to focus on parental perspectives rather than clinical measures, reduced the ability to assess the SOHPP’s impact. Finally, referring to all participants as “parents” for simplicity and consistency with the study aims, given that most (89%) identified as such, may have resulted in some overgeneralization.

Future research could address these gaps through several strategies. Longitudinal studies tracking children’s oral health behaviors and clinical outcomes before and after SOHPP implementation would enable causal inferences and quantify program impact. These stuies should incorporate objective measures, such as dental examinations or behavioral logs completed by children or teachers, alongside parental reports. Including rural and semi-urban populations in sampling frames would enhance generalizability. Qualitative studies, such as focus groups or in-depth interviews with parents, could explore cultural and systemic barriers to SOHPP uptake in-depth. Additionally, the roles of household head occupation and household size in shaping parental perceptions of the program could be further explored.

## Conclusions

This study provides a comprehensive assessment of parental perceptions within a sample participating in Iran’s SOHPP, highlighting both strengths and areas for improvement. Parents were generally aware of and satisfied with the program, especially its educational and preventive components, accepted fluoride therapy for their child, and perceived improvements in their child’s tooth brushing. However, they noted uneven changes in actual oral health behaviors, revealing a perception–behavior gap. Perceptions also varied by sociodemographic factors: more educated fathers reported greater awareness, satisfaction, and fluoride acceptance, while government-employed household heads were less aware and less likely to perceive positive effects on tooth-brushing. To enhance parental perceptions and long-term program impact, efforts should focus on strengthening treatment-related components and improving communication and procedural transparency around fluoride therapy. Simultenously, supporting consistent tooth-brushing and healthier snacking among children alongside tailoring outreach to parental sociodemographic characteristics is recommended.

## Data Availability

All relevant data are within the manuscript and its Supporting Information files.

## Acknowledgements

We would like to wholeheartedly thank all those who contributed to this study, particularly Dr. Mohammad Javad Kharazi Fard for his assistance with statistical analysis. We would also like to acknowledge the use of GPT-5 for editing this manuscript’s text, with prompts to improve text conciseness, readability, and coherence.

## Supporting information

**S1 file: English translation of the study questionnaire. S2 file: Anonymized dataset.**

